# Early Biological Markers of Post-Acute Sequelae of SARS-CoV-2 Infection

**DOI:** 10.1101/2023.07.14.23292649

**Authors:** Scott Lu, Michael J. Peluso, David V. Glidden, Michelle C. Davidson, Kara Lugtu, Jesus Pineda-Ramirez, Michel Tassetto, Miguel Garcia-Knight, Amethyst Zhang, Sarah A. Goldberg, Jessica Y. Chen, Maya Fortes-Cobby, Sara Park, Ana Martinez, Matthew So, Aidan Donovan, Badri Viswanathan, Rebecca Hoh, Kevin Donohue, David R. McIlwain, Brice Gaudiliere, Khamal Anglin, Brandon C. Yee, Ahmed Chenna, John W. Winslow, Christos Petropoulos, Steven G. Deeks, Melissa Briggs-Hagen, Raul Andino, Claire M. Midgley, Jeffrey N. Martin, Sharon Saydah, J. Daniel Kelly

**Author notes:** Indicates equal contribution (Scott Lu, Michael J. Peluso, Sharon Saydah, J. Daniel Kelly). “The findings and conclusions in this report are those of the authors and do not necessarily represent the official position of the Centers for Disease Control and Prevention”.

## Abstract

To understand the roles of acute phase viral dynamics and host immune responses in PASC, we enrolled 136 participants within 5 days of their first positive SARS-CoV-2 real-time PCR. Participants self-collected nasal specimens up to 21 times within the first 28 days after symptom onset; Interviewer-administered clinical questionnaires and blood samples were collected at enrollment and days 9, 14, 21, 28, and month 4 and 8 post-symptom. Defining PASC as the presence of any symptom new or worse since infection reported at their 4-month visit, we compared viral markers (quantity and duration of viral RNA load, infectious viral load, and plasma N-antigen level) and host immune markers (IL-6, IL-10, TNF-α, IFN-α, IFN-γ, MCP, IP-10, and Spike IgG) over the acute period. In comparison to those who fully recovered, those who developed PASC demonstrated significantly higher maximum levels of SARS-CoV-2 RNA, infectious virus, and N-antigen, longer duration of viral shedding, and lower Spike-specific IgG levels within the first 10 days of the acute phase of illness. No significant differences were identified among a panel of host immune markers, though there was a trend toward higher initial levels of certain markers (e.g., MCP-1, IFN-α, and IFN-γ) in those who went on to develop PASC. Early viral dynamics and the associated host immune responses play a role in the pathogenesis of PASC. These findings highlight the importance of understanding the early biological markers from acute SARS-CoV-2 infection in the natural history of PASC.

**Onset Sentence Summary:** Early viral dynamics and the associated host immune responses play a role in the pathogenesis of PASC.

## Introduction

Post-acute sequelae of SARS-CoV-2 infection (PASC) includes ongoing symptoms in the months following acute SARS-CoV-2 infection. Estimates of the occurrence of PASC can vary, currently approximately 1 in 10 U.S. adults who have had COVID-19 reporting ongoing symptoms (1). Despite a growing understanding of its epidemiology and natural history, the pathogenesis of PASC remains incompletely understood (2). Multiple mechanisms that could contribute to this condition are now under investigation (2, 3).

Viral antigen persistence and immune dysregulation are two mechanisms that might drive PASC (4–9). For example, recent work has suggested that a high proportion of individuals with PASC demonstrate detectable SARS-CoV-2 antigen in blood plasma during the post-acute phase (4, 6), and subgenomic RNA has been identified in widespread tissue sites at autopsy for up to 6 months post-COVID (5). Furthermore, studies comparing individuals with PASC with those who report complete recovery from SARS-CoV-2 infection have demonstrated increased levels of certain inflammatory markers including IL-6, TNF-alpha, and IL-1B, among others, for at least a year following infection (7–9). However, most work on PASC to date has been limited to the study of biological samples collected during the post-acute phase of infection.

Biological samples for predictors of PASC from the acute and early post-acute phase are more limited. Most studies from early infection have focused on shorter-term outcomes in hospitalized individuals (10, 11), although more recently some studies have suggested that prolonged viral clearance (12) or distinct early immune signatures (13) could be associated with PASC. While informative, these studies have been limited by relatively small study populations and/or less frequent biospecimen collection timepoints.

Here, we leveraged a household-based cohort of individuals intensively sampled during the acute phase of SARS-CoV-2 infection to assess the early biological determinants of PASC. We hypothesized that selected biological marker activity during early infection play a role in the pathogenesis of persistent symptomatology following the acute period of illness.

## Methods

### Study Population and Procedures

This was a longitudinal cohort study enrolling individuals acutely infected with SARS-CoV-2 and their household contacts in the San Francisco Bay area between September 2020 and May 2022. Details on the protocol and procedures for enrollment have been described in detail elsewhere (14). In brief, we enrolled participants who were recently diagnosed with nucleic acid-confirmed SARS-CoV-2 infection within 5 days of symptom onset. Molecular testing results from UCSF-affiliated sites were used by research staff to screen and recruit participants via in-person and telephone interviews. A study field team visited participants in their areas of isolation a total of 5 times during acute infection at the following timepoints: day of enrollment (ranging between day 0 and day 5 following self-reported symptom onset), and day 9, 14, 21, and 28 following symptom onset. At each study visit, blood and nasal specimens were collected. In addition, participants self-collected swabs of the anterior nares for the first 14 days post-infection, and at d17, 19, 21, and 28. Research coordinators administered phone interviews on the same day as the field visit within the acute period, then at 4 and 8 (+/- 2 months) months post-infection, using identical instruments developed in conjunction with a study of the post-acute phase (15).

### Measurements

#### Survey-based

We piloted a questionnaire containing items with key sociodemographic, medical history, symptomatology, quality-of-life, and later vaccination information. This instrument was developed with infectious disease specialists and epidemiologists who had managed COVID-19 patients. The form was iteratively developed over time and continues to be developed during the present study. Symptom items included a checklist of 32 selected symptoms followed by a free response option. The surveys are designed to assess symptom status from the time of the last interview to the current one. All symptoms reported as new, worsened or persistent since SARS-CoV-2 infection were captured; presence of symptoms prior to acute COVID-19 illness were assessed for exclusion from later analyses. Comorbid conditions include history of autoimmune disease, cancer, diabetes, HIV/AIDS, heart disease, hypertension, lung disease, and kidney disease.

The primary outcome of PASC was defined by the presence of any symptom reported between 2 to 6 months after initial illness. For participants whose first follow-up visit occurred beyond 6 months after initial illness, we analyzed symptom presence within the 2-6m PASC ascertainment window.

Information on the type and number of SARS-CoV-2 vaccinations were collected at baseline and updated with each interview. Fully vaccinated participants were defined as having received a complete primary vaccine series at least 14 days prior to study enrollment.

#### Virology

Details of sample collection, RNA extraction, and assay methodology have been reported previously (16). In short, acute viral RNA levels in nasal specimens were assessed using quantitative real time reverse transcription polymerase chain reaction (PCR) using the KingFisher platform to target nucleocapsid (N) and envelope (E) gene regions. A SARS-CoV-2 RNA-positive result was defined as having any level of RNA-positivity in N and E gene regions. Presence of infectious virus (infectivity) was measured via cytopathic effect (CPE) in Vero-hACE2-TMPRSS2 cells. Infectious viral titers were assessed using conventional plaque assay. All samples were tested by the same two researchers (MGK and MT).

Collected blood specimens were also used to measure nucleocapsid antigen (N-Ag) at all available time points.

#### Host immune factors

Plasma biomarker measurements were performed using the automated HD-X Simoa platform. Analytes included plasma SARS-CoV-2 Spike receptor binding domain (RBD) IgG and the following inflammatory markers: IL-6, IL-10, TNF-α, IFN-α, IFN-γ, IP-10, and MCP-1.

All assays were performed according to the manufacturer’s instructions and assay performance was consistent with the manufacturer’s specifications.

### Statistical Analysis

We describe distribution of virologic and host immune factors at baseline and through follow-up among those with PASC compared to those without PASC. Virologic factors included the following:

- RNA viral load over time
- Maximum RNA load
- Duration of RNA detection (defined as days from illness onset to last positive PCR)
- RNA decay rates (defined as linear rate of RNA decline from maximum RNA load)
- Infectious viral titer on day of max RNA
- Duration of detection of culturable virus (defined at days from illness onset to last positive CPE)
- N antigen levels
- N antigen decay rates (defined as linear rate of N antigen decline).

Non-infectious and infectious viral shedding and viral load from PCR of nasal specimens were compared using logistic regression followed by marginal effects to calculate adjusted risk ratios (aRR) (17). Subgroup analyses were conducted on participants with available blood specimens and included N antigen and host immune factors. Missing data among the subgroup was assessed and managed using multivariate imputation by chained equations (18). After imputation, we fit linear models using generalized estimating equations with a working independence correlation matrix, identity linkage, and Gaussian distribution using terms for PASC, time period, and interaction between them. The distribution and magnitude of difference of each analyte are summarized graphically.

Given the nonlinear presentation of viral load and antibody results we opted to log transform results for comparison. For antibody analysis we further restricted our model to unvaccinated participants in order to compare antibody responses. Study data was captured using REDCap electronic data capture tools. All estimates were calculated using Stata (version 16.1; StataCorp, College Station, TX).

### Human Subjects

The study was reviewed by the University of California, San Francisco (UCSF) Institutional Review Board and given a designation of public health surveillance according to federal regulations as summarized in 45 CFR 46.102(d)(1)(2). Informed consent was obtained from all participants. This activity was reviewed by CDC and was conducted consistent with applicable federal law and CDC policy*

## Results

### Study participants

From September 2020 to May 2022, we enrolled 136 SARS-CoV-2 infected participants during the acute phase of their illness. Among these participants, 104 completed at least one post-acute visit and contributed at least one nasal sample for infectious viral and RNA testing (87% of participants with 10 or more nasal specimens). These participants were all enrolled during their first confirmed case of COVID-19. Among these 104, 80 participants also contributed blood for viral N-antigen and inflammatory marker testing.

Participants had a median age of 35.5 years (IQR: 27 to 44), and the cohort was diverse in terms of sex and race and ethnicity (Table 1). Participants were generally healthy and most (77%) did not report pre-existing comorbid medical conditions; the most common comorbidities were lung disease (14%), hypertension (10%), and diabetes (5%). The majority of participants (93%) were infected with pre-Omicron strains of SARS-CoV-2 and most (65%) had not received a SARS-CoV-2 vaccine prior to their infection. Over the course of their acute illness, 96 of 104 (92%) participants reported presence of at least one symptom with a median of 9 symptoms (IQR: 4 to 13). The most common acute symptoms were fatigue (76%), cough (71%), rhinorrhea (71%), headache (54%), and sore throat (46%). See Supplemental Table 1 for characteristics of the subgroup of participants who had viral N-antigen and cytokine testing. Participants had similar characteristics as those in the overall cohort, except that all participants in the subgroup were infected with a pre-Omicron variant.

**Table 1.**
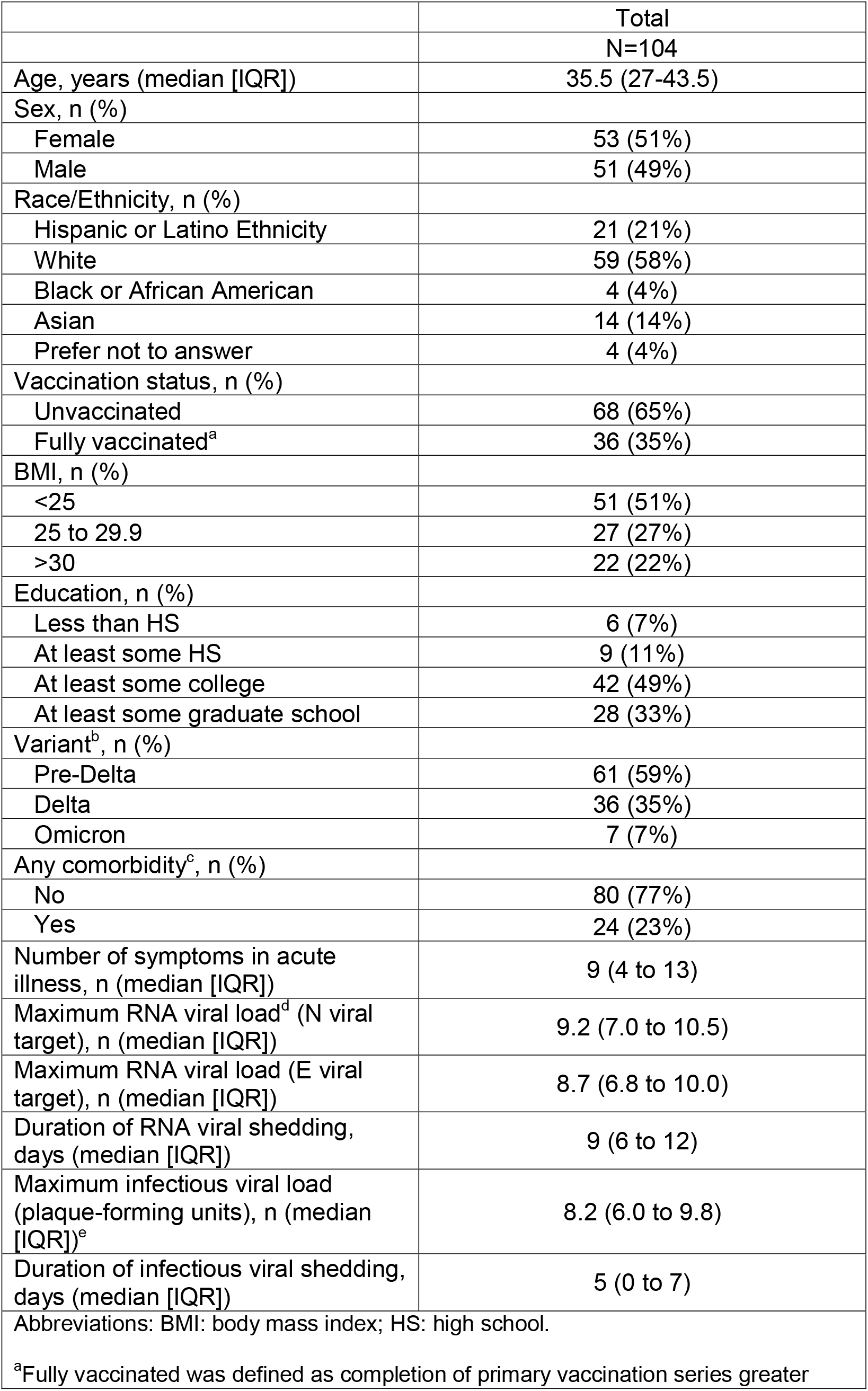
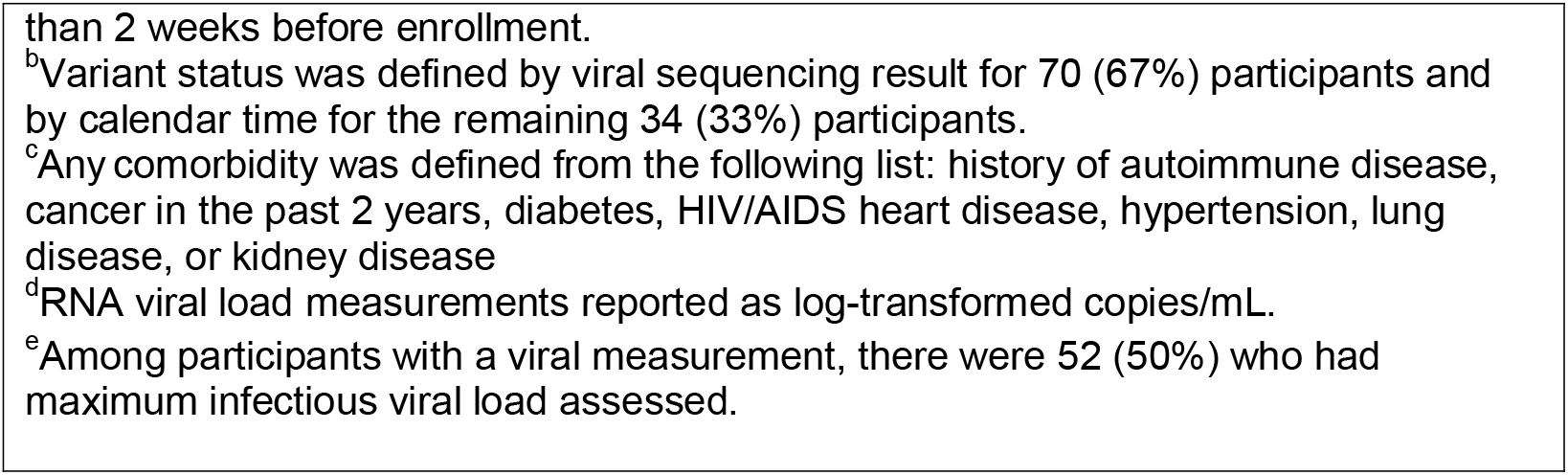
Baseline characteristics of the cohort.

|  | Total |
| --- | --- |
|  | N=104 |
| Age, years (median [IQR]) | 35.5 (27-43.5) |
| Sex, n (%) |  |
| Female | 53 (51%) |
| Male | 51 (49%) |
| Race/Ethnicity, n (%) |  |
| Hispanic or Latino Ethnicity | 21 (21%) |
| White | 59 (58%) |
| Black or African American | 4 (4%) |
| Asian | 14 (14%) |
| Prefer not to answer | 4 (4%) |
| Vaccination status, n (%) |  |
| Unvaccinated | 68 (65%) |
| Fully vaccinated<sup>a</sup> | 36 (35%) |
| BMI, n (%) |  |
| <25 | 51 (51%) |
| 25 to 29.9 | 27 (27%) |
| >30 | 22 (22%) |
| Education, n (%) |  |
| Less than HS | 6 (7%) |
| At least some HS | 9 (11%) |
| At least some college | 42 (49%) |
| At least some graduate school | 28 (33%) |
| Variant<sup>b</sup>, n (%) |  |
| Pre-Delta | 61 (59%) |
| Delta | 36 (35%) |
| Omicron | 7 (7%) |
| Any comorbidity<sup>c</sup>, n (%) |  |
| No | 80 (77%) |
| Yes | 24 (23%) |
| Number of symptoms in acute illness, n (median [IQR]) | 9 (4 to 13) |
| Maximum RNA viral load<sup>d</sup> (N viral target), n (median [IQR]) | 9.2 (7.0 to 10.5) |
| Maximum RNA viral load (E viral target), n (median [IQR]) | 8.7 (6.8 to 10.0) |
| Duration of RNA viral shedding, days (median [IQR]) | 9 (6 to 12) |
| Maximum infectious viral load (plaque-forming units), n (median [IQR])<sup>e</sup> | 8.2 (6.0 to 9.8) |
| Duration of infectious viral shedding, days (median [IQR]) | 5 (0 to 7) |
| Abbreviations: BMI: body mass index; HS: high school. |  |
| <sup>a</sup>Fully vaccinated was defined as completion of primary vaccination series greater |  |

than 2 weeks before enrollment.
<sup>b</sup>Variant status was defined by viral sequencing result for 70 (67%) participants and by calendar time for the remaining 34 (33%) participants.
<sup>c</sup>Any comorbidity was defined from the following list: history of autoimmune disease, cancer in the past 2 years, diabetes, HIV/AIDS heart disease, hypertension, lung disease, or kidney disease
<sup>d</sup>RNA viral load measurements reported as log-transformed copies/mL.
<sup>e</sup>Among participants with a viral measurement, there were 52 (50%) who had maximum infectious viral load assessed.

### Description of PASC

Among 104 SARS-CoV-2 infected participants with a post-acute follow-up visit, 31 (30%) reported new, worsened or persistent symptoms since the time of SARS-CoV-2 infection, consistent with PASC. We did not identify a difference between those with and without PASC in terms of vaccination status (31% vs 36%). All participants with PASC had been symptomatic during acute illness with a median of 12 (9 to 14); among those without PASC the median number of symptoms in the acute period was 7 (3 to 12).

Those meeting criteria for PASC reported a median of 2 (IQR: 1 to 6) symptoms during the post-acute period 2-6 months after SARS-CoV-2 infection. The most commonly reported PASC symptoms were fatigue (32%), difficulty with concentration/memory (33%), rhinorrhea (29%), cough (29%), and headache (26%).

### Associations of virologic factors with PASC

We compared the magnitude, decay rates and duration of RNA and infectious viral shedding and N-antigen levels between participants who did and did not develop PASC (**Table 2**). Compared to those who did not develop PASC, those who developed PASC had higher maximum RNA viral load, infectious viral load, and N-antigen levels (**Table 2**; **Figure 2a**). Viral RNA decay rates did not significantly differ by PASC status, but decay rates of N-antigen levels were faster in the first 9 days after symptom onset among those with PASC than those without PASC (**Figure 2a**).

**Table 2.** Association of virologic factors with post-acute sequelae of SARS-CoV-2 infection (PASC) (N=104).

|  | Non-PASC<br>N=73 | PASC<br>N=31 | Adjusted Risk Ratios<sup>c</sup><br>(95% CI) |
| --- | --- | --- | --- |
| Maximum RNA N viral load<sup>a</sup>, n<br>(median [IQR]) | 8.4 (6.7 to 10.1) | 10.0 (8.9 to 10.9) | 1.30 (1.03 to 1.66) |
| Maximum RNA E viral load, n<br>(median [IQR]) | 8.2 (6.6 to 9.8) | 9.1 (8.4 to 10.4) | 1.26 (0.99 to 1.61) |
| Rate of RNA N viral decay | -0.3 (-0.6 to -0.1) | -0.4 (-0.5 to -0.2) | 0.59 (0.18 to 1.96) |
| Rate of RNA E viral decay | -0.4 (-0.6 to -0.2) | -0.5 (-0.7 to -0.3) | 0.69 (0.31 to 1.52) |
| Duration of RNA viral<br>shedding, days (median [IQR]) | 9 (6 to 11) | 10.5 (8 to 13) | 1.47 (0.80 to 2.70) |
| Maximum infectious viral load,<br>n (median [IQR])<sup>b</sup> | 6.9 (5.4 to 9.5) | 9.0 (8.4 to 10.3) | 1.35 (1.07 to 1.70) |
| Duration of infectious viral<br>shedding, days (median [IQR]) | 5 (0 to 7) | 6 (2 to 8) | 1.08 (0.98 to 1.20) |
| <sup>a</sup>RNA viral load measurements were log-transformed copies/mL. |  |  |  |
| <sup>b</sup>Among 104 participants with a viral measurement, there were 94 (90%) who had maximum infectious viral load assessed. |  |  |  |
| <sup>c</sup>Adjusted risk ratios calculated with delta-method standard errors. |  |  |  |

**Figure 1.**
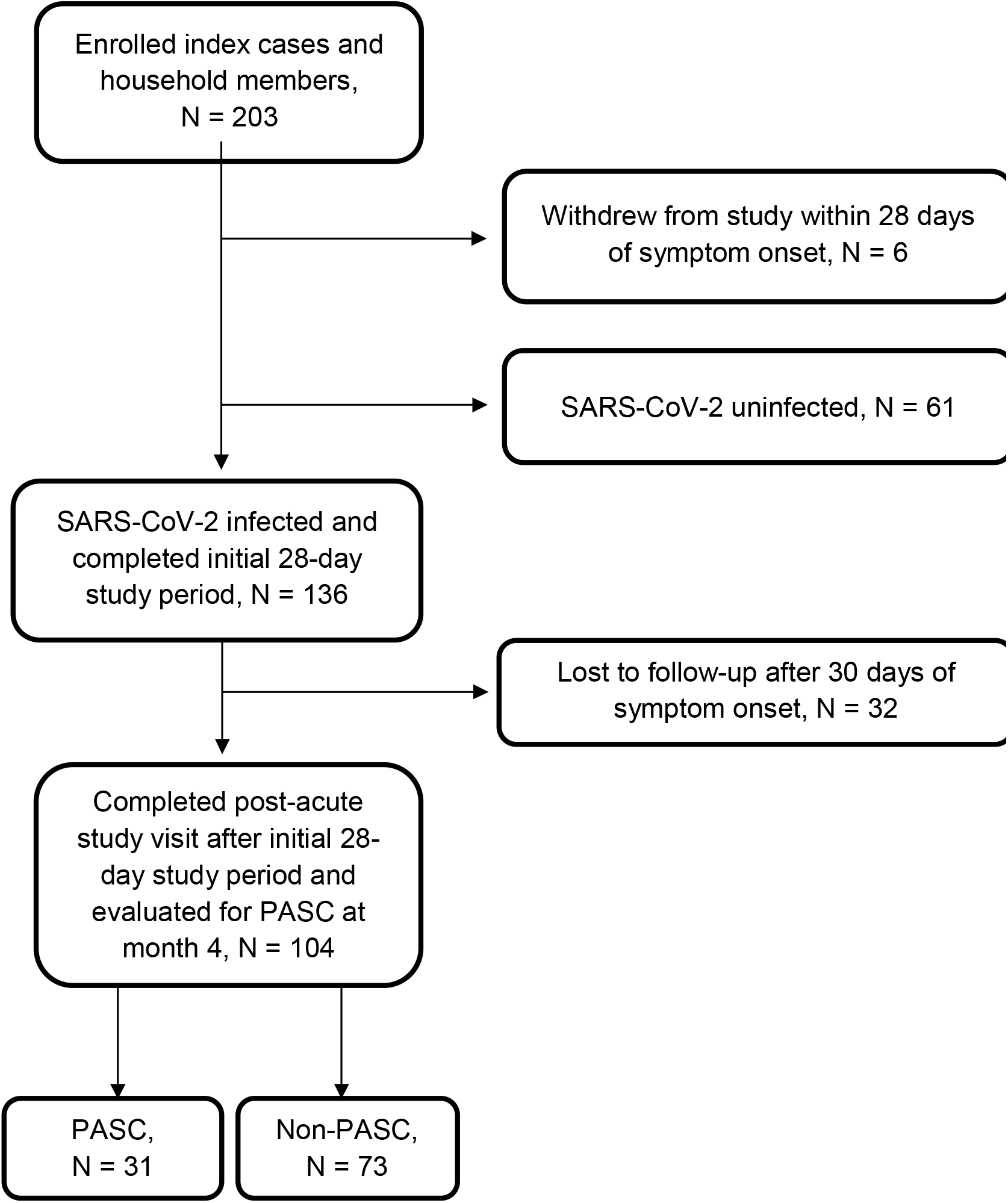
Flow diagram of participants evaluated for post-acute sequelae of SARS-CoV-2 infection (PASC) at month 4.

**Figure 2.**
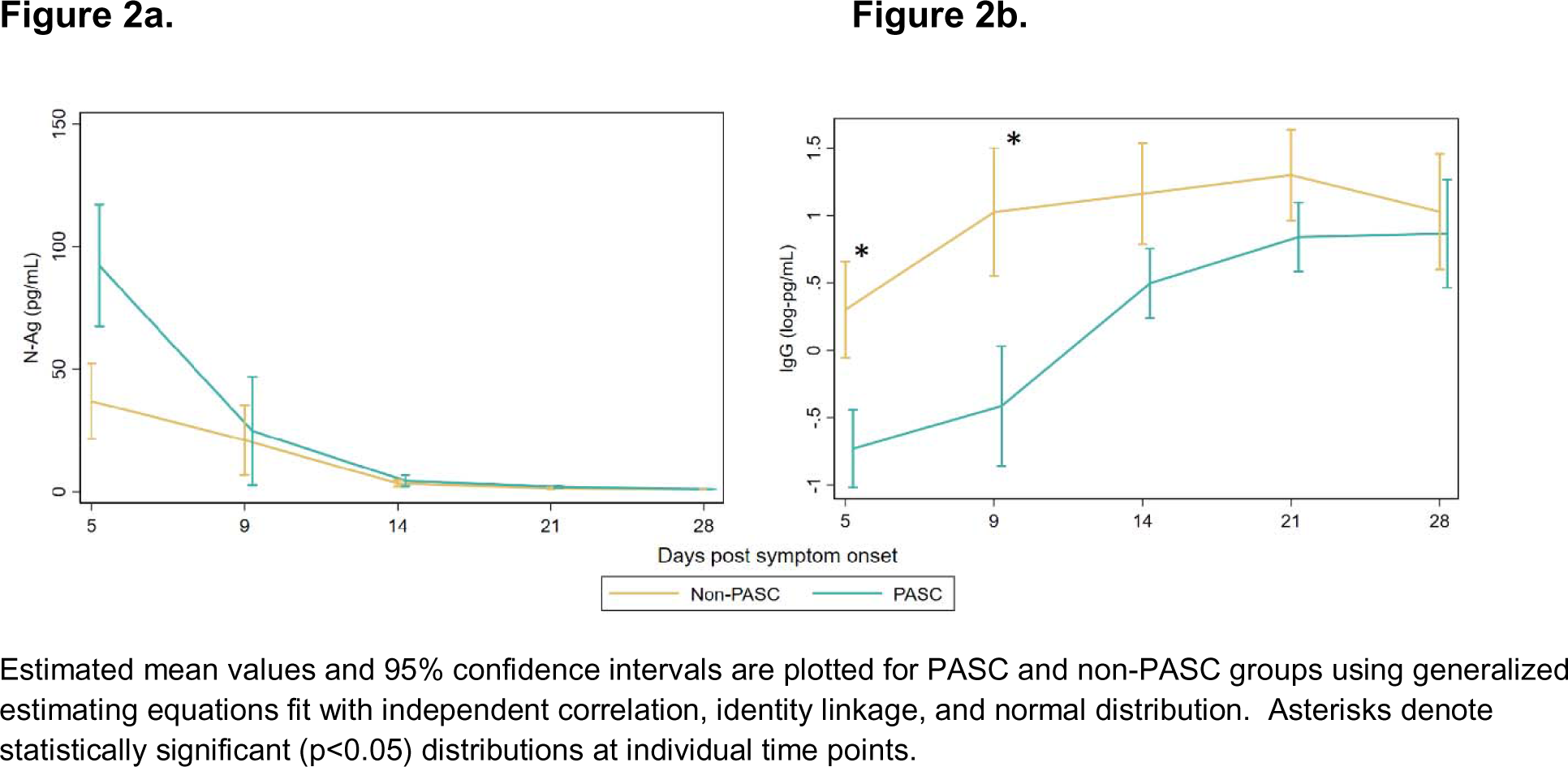
Comparison of N-antigen and IgG Spike antibody levels among those with and without post-acute sequelae of SARS-CoV-2 (PASC) over a 28-day period after symptom onset. Figure 2a shows N antigen levels. Figure 2b shows IgG Spike antibody levels among unvaccinated participants.

The development of PASC was associated with a higher proportion of RNA positivity (p = 0.01) and infectious viral shedding (p = 0.02; Figure 3). Furthermore, participants who went on to develop PASC had a higher median number of days of RNA and infectious viral shedding compared with those who did not develop PASC, though these differences did not reach statistical significance (**Table 2**).

**Figure 3.**
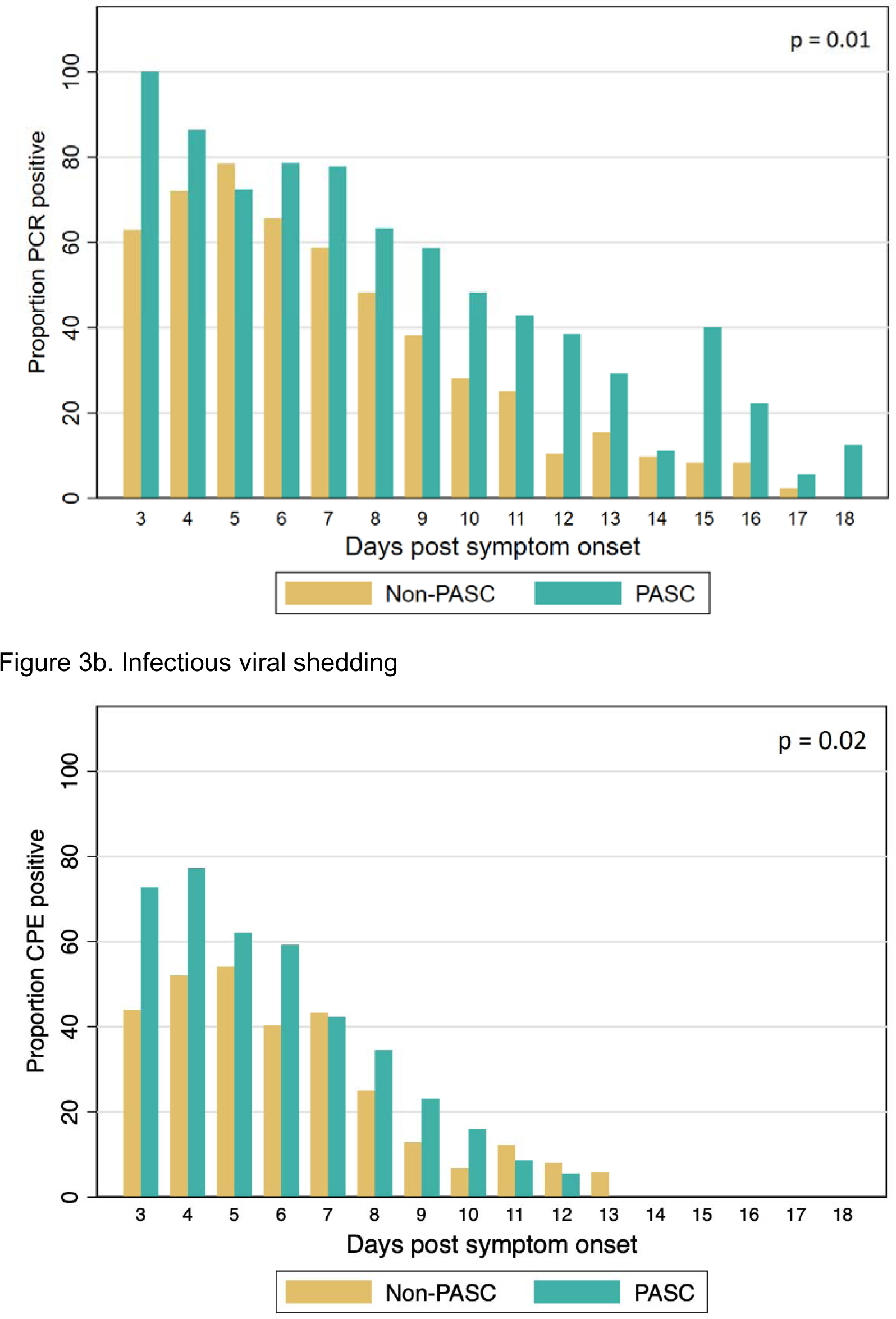
Proportion of RNA and infectious viral shedding for each day after symptom onset among those with and without PASC. Figure 3a depicts RNA viral shedding while Figure 3b describes infectious viral shedding over an 18-day period.

### Associations of immune responses with PASC

As expected, anti-RBD IgG levels among unvaccinated participants climbed over the 28-day observation period. Notably, those who developed PASC exhibited lower levels of Spike IgG at day 5 (p=0.03) and day 9 (p=0.03) post-symptom onset compared to those who did not develop PASC (**Figure 2b**). These differences attenuated after day 14 and both groups had similar Spike IgG levels by day 28.

The trajectories of inflammatory biomarkers markers followed expected declining trends during the first 28 days following symptom onset (**Figure 4**). Those who developed PASC had non-significant trends toward higher initial levels of MCP, IFN-α, and IFN-γ that attenuated over the observation period. Overall, there were no strong associations between starting levels or trajectories of inflammatory markers during the acute phase and the later development of PASC.

**Figure 4.**
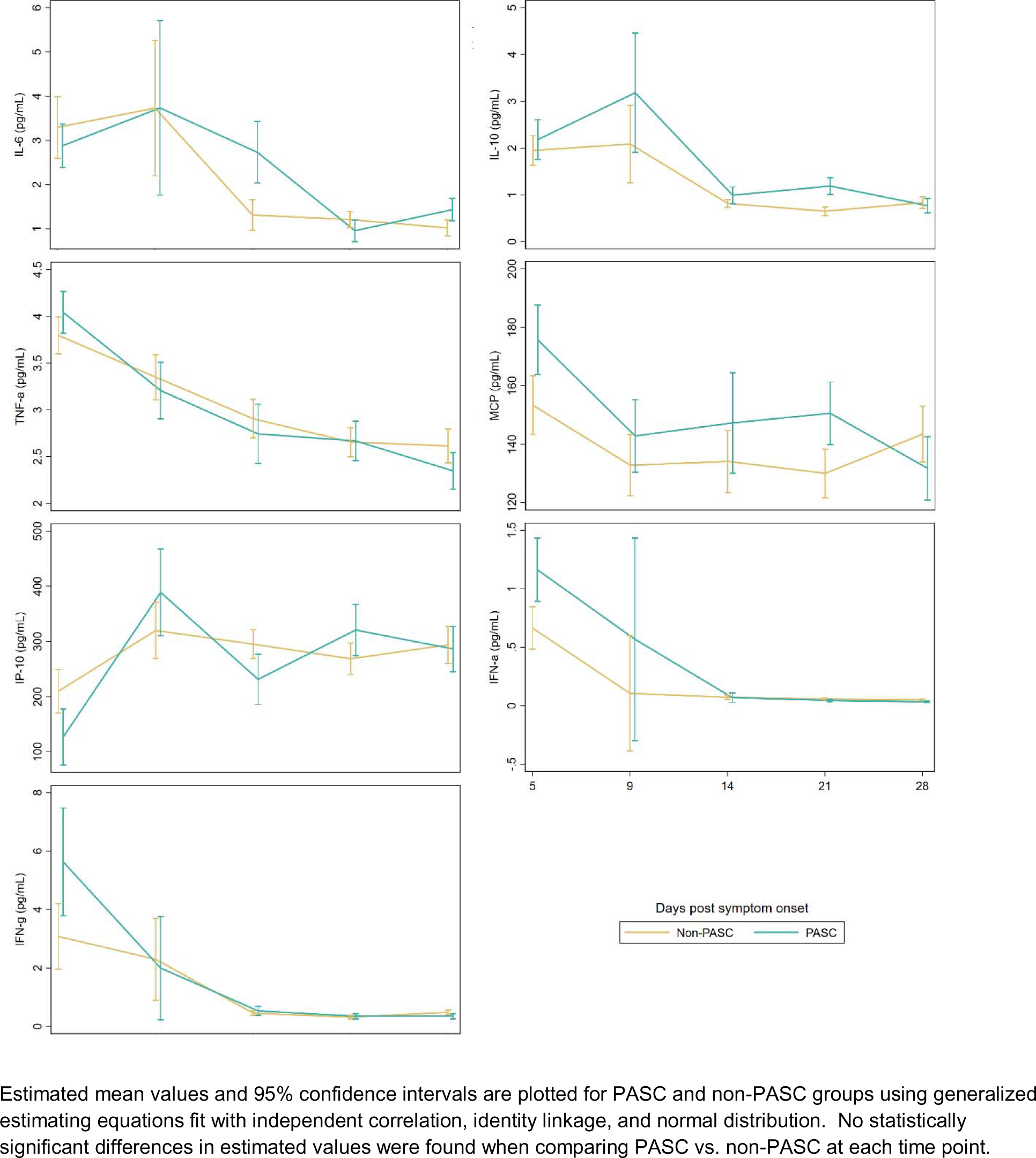
Inflammatory marker levels among those with and without post-acute sequelae of SARS-CoV-2 infection (PASC) over a 28-day period after symptom onset.

## Discussion

In this household-based cohort of outpatients followed prospectively from the time of SARS-CoV-2 symptom onset through the post-acute period, we identified several early biologic determinants of PASC. Specifically, we found a relationship between early viral dynamics, adaptive immune responses, and the later development of PASC. These findings suggest that an individual’s ability to control viral replication and ongoing viral persistence may play an important role in recovery from SARS-CoV-2 infection, and that the dynamics of the immune responses during the acute and early post-acute phase may determine who goes on to experience post-acute symptoms. Our observations also provide additional rationale for the evaluation of early antiviral therapy as one potential target for mitigating the development of PASC.

The etiology of PASC remains incompletely understood and there are no specific treatments available beyond symptom management. To date, most biological assessments of PASC have focused on the post-acute phase of infection (4, 6–9, 19–25). Only a few studies have assessed the relationship between biomarkers in the acute phase of SARS-CoV-2 infection and the later development of PASC (12, 13, 26). Such efforts have identified the presence of SARS-CoV-2 RNA in blood at the time of diagnosis (26) and prolonged duration of viral shedding from the upper respiratory tract (12) as correlates of PASC. Overall, these efforts to understand the relationship between acute phase biology and post-acute symptomatology have been limited by the considerable challenges related to collecting specimens during the earliest days following COVID-19 symptom onset, and as a result many studies have been limited to individuals hospitalized with COVID-19, who are not representative of most individuals experiencing PASC.

Using orthogonal virologic assessments, we observed a relationship between the early viral burden and the development of PASC. Those who went on to develop PASC had higher maximum levels of SARS-CoV-2 RNA, infectious virus, and viral N-antigen protein during acute infection, and demonstrated more rapid decay of N-antigen levels during the first 9 days after symptom onset, suggesting that either higher antigen burden or a more robust immune response in response to that burden are related to the later development of PASC. Furthermore, we found that those who later developed PASC exhibited more prolonged shedding of infectious virus and had a less robust humoral immune response in the first 9 days following infection, potentially related to more sluggish viral clearance. Taken together, these findings suggest that the total amount of virus present, the immunologic response that results in control of that virus, and the efficiency of clearing infectious virus might all drive the development of post-acute symptoms.

A number of demographic and clinical risk factors for PASC have been identified. While prior SARS-CoV-2 vaccination appears to be protective against the development of PASC (27), it remains unclear whether antiviral treatment during the acute phase of illness might reduce the incidence of this condition. Two studies suggest a benefit among those meeting criteria for antiviral therapy (28, 29), but other evaluations have not shown an effect (30, 31). Our findings suggest that interventions that alter the magnitude or duration of infectious virus and the associated immune response during the acute phase might have the potential to affect the development of PASC. These might include antivirals (e.g., protease or RNA polymerase inhibitors) to reduce viral replication and/or monoclonal antibodies or therapeutic vaccination to enhance the immune response needed to neutralize the virus. Assessment of the impact of these interventions during the acute phase of infection is warranted, although we note that most of the cohort included in this analysis was enrolled prior to the availability of SARS-CoV-2 vaccines and that preexisting immunity related to prior vaccination or infection might alter these mechanisms.

Acute COVID-19 is a highly inflammatory condition (10, 11). While prior cross-sectional and longitudinal analyses in the post-acute phase have demonstrated that PASC is associated with differences in markers of immune activation (7–9, 19, 21, 26), we did not identify significant differences in levels of these markers during the acute phase between those who did and did not go on to develop PASC. There are several possible explanations for this. First, our cohort was likely to be more homogeneous in terms of disease severity, with a lower proportion of those with severe enough acute illness to be unable to complete the study’s arduous questionnaire and specimen collection schedule and none who were hospitalized, than those included in other studies. Second, the number of individuals developing PASC was relatively small, limiting our statistical power to observe clear differences even when trends were present. Third, it is possible that these differences develop over time during the post-acute phase, in response to ongoing immunologic activity, and for that reason may not be present during the acute and early post-acute timepoints in this analysis.

Strengths of our study include the high degree of adherence to the biospecimen collection protocol and the collection of biospecimens in close proximity to initial symptom onset in a cohort of outpatients, who comprise the vast majority of individuals with PASC. The inclusion of a large proportion of individuals from earlier waves of the pandemic, prior to the widespread availability of vaccination, SARS-CoV-2 treatment, or reinfection allowed us to study the natural history of this condition in the absence of these confounding factors. However, this analysis has several limitations. First, due to relatively small cohort size, we were unable to evaluate whether early markers were associated with specific symptomatic phenotypes of PASC, each of which may be driven by different biology. Second, we defined PASC as the presence of any symptom new or worse since SARS-CoV-2 infection present during the post-acute period, without requiring that the same symptom be present during the acute period; it is possible that this could result in misattribution of some unrelated symptoms to SARS-CoV-2 infection. This is mitigated somewhat by specific training for study staff to interrogate for presence of any reported symptom prior to SARS-CoV-2 infection. Third, we applied a relatively broad time window in which PASC could be defined (2 to 6 months), which although consistent with accepted case definitions might be subject to variability by time since infection (either waxing and waning or resolving symptoms). Future assessment of individuals during later timepoints (e.g., 6 months or beyond) following infection may be warranted. Fourth, the current analysis focuses on the binary presence or absence of symptoms and does not include consideration of symptom severity or impact. Such assessment could lead to further insight into whether different viral titers and immune responses can impact the severity of symptoms rather than simply their presence or absence. Finally, although mitigated through use of multiple imputation, missing data from declined blood sample collection could be a source of error.

In summary, viral load and timing of antibody development may be important factors in the pathogenesis of PASC. These early biological markers may be part of a larger cascade of events during the earliest days of SARS-CoV-2 infection that warrant consideration in the larger efforts to develop a mechanistic understanding of this condition. Further research during the acute phase of SARS-CoV-2 infection is required to elucidate causal mechanisms of PASC, which can eventually lead to the development of therapeutics to prevent or treat this condition.

## Data Availability

All data produced in this present work are contained within the manuscript and available upon reasonable request to the authors.

## Acknowledgements

We are grateful to the study participants. We would like to acknowledge Jeremy Lambert and Quanterix Inc for providing SARS-CoV2 N-antigen and anti-RBD IgG assay kits.

## Author contributions

SL, MJP, JDK, SS, and KNM designed the study supported by funding to JDK. SL, MCD, KL, JPR, SAG, JYC, MFC, SP, AM, KA, and JDK collected clinical data and biospecimens. MT, MGK, AZ, DRM, BG, BCY, AC, JWW, and CP were responsible for biospecimen processing and laboratory testing. SL,DVG, and JDK performed and/or interpreted the statistical analysis. SL, MJP, and JDK drafted the initial manuscript and edited the

## Data and Material Availability

All data associated with this study are present in the paper. Patient data can only be shared upon request to JDK.

## Supplementary Material

**Supplemental Table 1:** Participant characteristics by outcome status.

|  | Non-PASC | PASC | Total |
| --- | --- | --- | --- |
|  | N=73 | N=31 | N=104 |
| Age, years (median [IQR]) | 32 (24-41) | 39 (32-51) | 35.5 (27-43.5) |
| Sex, n (%) |  |  |  |
| Female | 37 (51%) | 16 (52%) | 53 (51%) |
| Male | 36 (49%) | 15 (48%) | 51 (49%) |
| Race/Ethnicity, n (%) |  |  |  |
| Hispanic or Latino Ethnicity | 16 (23%) | 5 (16%) | 21 (21%) |
| White | 40 (56%) | 19 (61%) | 59 (58%) |
| Black or African American | 3 (4%) | 1 (3%) | 4 (4%) |
| Asian | 10 (14%) | 4 (13%) | 14 (14%) |
| Prefer not to answer | 2 (3%) | 2 (6%) | 4 (4%) |
| Vaccination status, n (%) |  |  |  |
| Unvaccinated | 47 (64%) | 21 (68%) | 68 (65%) |
| Fully vaccinated<sup>a</sup> | 26 (36%) | 10 (32%) | 36 (35%) |
| BMI, n (%) |  |  |  |
| <25 kg/m<sup>2</sup> | 40 (58%) | 11 (35%) | 51 (51%) |
| 25 to 29.9 | 20 (29%) | 7 (23%) | 27 (27%) |
| >30 | 9 (13%) | 13 (42%) | 22 (22%) |
| Education, n (%) |  |  |  |
| Less than HS | 4 (7%) | 2 (7%) | 6 (7%) |
| At least some HS | 6 (11%) | 3 (10%) | 9 (11%) |
| At least some college | 29 (53%) | 13 (43%) | 42 (49%) |
| At least some graduate school | 16 (29%) | 12 (40%) | 28 (33%) |
| Variant<sup>b</sup>, n (%) |  |  |  |
| Pre-Delta | 41 (56%) | 20 (65%) | 61 (59%) |
| Delta | 26 (36%) | 10 (32%) | 36 (35%) |
| Omicron | 6 (8%) | 1 (3%) | 7 (7%) |
| Any comorbidity<sup>c</sup>, n (%) |  |  |  |
| No | 58 (79%) | 22 (71%) | 80 (77%) |
| Yes | 15 (21%) | 9 (29%) | 24 (23%) |
| Number of symptoms in acute illness | 7 (3 to 12) | 12 (9 to 14) | 9 (4 to 13) |

Abbreviations: BMI: body mass index; HS: high school.
<sup>a</sup>Fully vaccinated was defined as completion of primary vaccination series with 2 week window prior to symptom onset.
<sup>b</sup>Variant status was defined by viral sequencing result for 70 (67%) participants and by calendar time for the remaining 34 (33%) participants.
<sup>c</sup>Any comorbidity was defined from the following list: history of autoimmune disease, cancer in the past 2 years, diabetes, heart disease, hypertension, lung disease, or kidney disease

**Supplemental Table 2:** Participant characteristics by outcome status, restricted to those who received a blood draw.

|  | No PASC | PASC | Total |
| --- | --- | --- | --- |
|  | N=52 | N=28 | N=80 |
| Age, years (median [IQR]) | 37 (29-44) | 39.5 (32.5-48.5) | 38 (30-45) |
| Sex, n (%) |  |  |  |
| Female | 28 (54) | 13 (46) | 41 (51) |
| Male | 24 (46) | 15 (54) | 39 (49) |
| Race/Ethnicity, n (%) |  |  |  |
| Hispanic or Latino Ethnicity | 10 (20) | 5 (18) | 15 (19) |
| White | 27 (54) | 17 (61) | 44 (56) |
| Black or African American | 2 (4) | 1 (4) | 3 (4) |
| Asian | 9 (18) | 3 (11) | 12 (15) |
| Prefer not to answer | 2 (4) | 2 (7) | 4 (5) |
| Vaccination status, n (%) |  |  |  |
| Unvaccinated | 30 (58) | 20 (71) | 50 (63) |
| Fully vaccinated<sup>a</sup> | 22 (42) | 8 (29) | 30 (38) |
| BMI, n (%) |  |  |  |
| <25 | 23 (47) | 11 (39) | 34 (44) |
| 25 to 29.9 | 18 (37) | 6 (21) | 24 (31) |
| >30 | 8 (16) | 11 (39) | 19 (25) |
| Education, n (%) |  |  |  |
| Less than HS | 3 (6) | 2 (7) | 5 (6) |
| At least some HS | 6 (12) | 3 (11) | 9 (12) |
| At least some college | 27 (53) | 11 (41) | 38 (49) |
| At least some graduate school | 15 (29) | 11 (41) | 26 (33) |
| Variant<sup>b</sup>, n (%) |  |  |  |
| Pre-Delta | 31 (60) | 19 (68) | 50 (63) |
| Delta | 21 (40) | 9 (32) | 30 (38) |
| Omicron | 0 (0) | 0 (0) | 0 (0) |
| Any comorbidity<sup>c</sup>, n (%) |  |  |  |
| No | 38 (73) | 20 (71) | 58 (73) |
| Yes | 14 (27) | 8 (29) | 22 (28) |
| Number of symptoms in acute illness | 9 (4.5 to 14) | 12 (10 to 14.5) | 11 (6 to 14) |
| Abbreviations: BMI: body mass index; HS: high school. |  |  |  |
| <sup>a</sup>Fully vaccinated was defined as completion of primary vaccination series with 2 week window prior to symptom onset. |  |  |  |
| <sup>b</sup>Variant status was defined by viral sequencing result for 70 (67%) participants and by calendar time for the remaining 34 (33%) participants. |  |  |  |
| <sup>c</sup>Any comorbidity was defined from the following list: history of autoimmune disease, cancer in the past 2 years, diabetes, heart disease, hypertension, lung disease, or kidney disease |  |  |  |

## Footnotes

* see e.g., 45 C.F.R. part 46.102(I)(2), 21 C.F.R. part 56; 42 U.S.C. §241(d); 5 U.S.C. §552a; 44 U.S.C. §3501 et seq..

## References and Notes

1. Bureau NCfHSUSC. Household Pulse Survey, 2022–2023. Long COVID. 2023 [Available from: https://www.cdc.gov/nchs/covid19/pulse/long-covid.htm.

2. Peluso MJ, Deeks SG. Early clues regarding the pathogenesis of long-COVID. Trends Immunol. 2022;43(4):268–70.

3. Davis HE, McCorkell L, Vogel JM, Topol EJ. Long COVID: major findings, mechanisms and recommendations. Nat Rev Microbiol. 2023:1–14.

4. Swank Z, Senussi Y, Manickas-Hill Z, Yu XG, Li JZ, Alter G, et al. Persistent circulating SARS-CoV-2 spike is associated with post-acute COVID-19 sequelae. Clin Infect Dis. 2022.

5. Stein SR, Ramelli SC, Grazioli A, Chung J-Y, Singh M, Yinda CK, et al. SARS-CoV-2 infection and persistence in the human body and brain at autopsy. Nature. 2022;612(7941):758–63.

6. Peluso MJ, Deeks SG, Mustapic M, Kapogiannis D, Henrich TJ, Lu S, et al. SARS-CoV-2 and Mitochondrial Proteins in Neural-Derived Exosomes of COVID-19. Ann Neurol. 2022.

7. Peluso MJ, Lu S, Tang AF, Durstenfeld MS, Ho H-E, Goldberg SA, et al. Markers of Immune Activation and Inflammation in Individuals With Postacute Sequelae of Severe Acute Respiratory Syndrome Coronavirus 2 Infection. J Infect Dis. 2021.

8. Phetsouphanh C, Darley DR, Wilson DB, Howe A, Munier CML, Patel SK, et al. Immunological dysfunction persists for 8 months following initial mild-to-moderate SARS-CoV-2 infection. Nat Immunol. 2022:1–7.

9. Schultheiß C, Willscher E, Paschold L, Gottschick C, Klee B, Henkes S-S, et al. The IL-1β, IL-6, and TNF cytokine triad is associated with post-acute sequelae of COVID-19. Cell Rep Med. 2022;3(6):100663.

10. Lucas C, Wong P, Klein J, Castro TBR, Silva J, Sundaram M, et al. Longitudinal analyses reveal immunological misfiring in severe COVID-19. Nature. 2020;584(7821):463–9.

11. Del Valle DM, Kim-Schulze S, Huang H-H, Beckmann ND, Nirenberg S, Wang B, et al. An inflammatory cytokine signature predicts COVID-19 severity and survival. Nat Med. 2020;26(10):1636–43.

12. Antar AAR, Yu T, Demko ZO, Hu C, Tornheim JA, Blair PW, et al. Long COVID brain fog and muscle pain are associated with longer time to clearance of SARS-CoV-2 RNA from the upper respiratory tract during acute infection. medRxiv. 2023.

13. Thompson RC, Simons NW, Wilkins L, Cheng E, Del Valle DM, Hoffman GE, et al. Molecular states during acute COVID-19 reveal distinct etiologies of long-term sequelae. Nat Med. 2022:1–11.

14. Kelly JD, Lu S, Anglin K, Garcia-Knight M, Pineda-Ramirez J, Goldberg SA, et al. Magnitude and Determinants of Severe Acute Respiratory Syndrome Coronavirus 2 (SARS-CoV-2) Household Transmission: A Longitudinal Cohort Study. Clin Infect Dis. 2022;75(Suppl 2):S193–S204.

15. Peluso MJ, Kelly JD, Lu S, Goldberg SA, Davidson MC, Mathur S, et al. Persistence, Magnitude, and Patterns of Postacute Symptoms and Quality of Life Following Onset of SARS-CoV-2 Infection: Cohort Description and Approaches for Measurement. Open Forum Infect Dis. 2022;9(2):ofab640.

16. Garcia-Knight M, Anglin K, Tassetto M, Lu S, Zhang A, Goldberg SA, et al. Infectious viral shedding of SARS-CoV-2 Delta following vaccination: A longitudinal cohort study. PLoS Pathog. 2022;18(9):e1010802.

17. Norton EC, Miller MM, Kleinman LC. Computing Adjusted Risk Ratios and Risk Differences in Stata. The Stata Journal. 2013;13(3):492–509.

18. White IR, Royston P, Wood AM. Multiple imputation using chained equations: Issues and guidance for practice. Statistics in Medicine. 2011;30(4):377–99.

19. Peluso MJ, Sans HM, Forman CA, Nylander AN, Ho H-E, Lu S, et al. Plasma Markers of Neurologic Injury and Inflammation in People With Self-Reported Neurologic Postacute Sequelae of SARS-CoV-2 Infection. Neurol Neuroimmunol Neuroinflamm. 2022;9(5).

20. Durstenfeld MS, Peluso MJ, Kelly JD, Win S, Swaminathan S, Li D, et al. Role of antibodies, inflammatory markers, and echocardiographic findings in postacute cardiopulmonary symptoms after SARS-CoV-2 infection. JCI Insight. 2022;7(10).

21. Klein J, Wood J, Jaycox J, Lu P, Dhodapkar RM, Gehlhausen JR, et al. Distinguishing features of Long COVID identified through immune profiling. medRxiv. 2022.

22. Son K, Jamil R, Chowdhury A, Mukherjee M, Venegas C, Miyasaki K, et al. Circulating anti-nuclear autoantibodies in COVID-19 survivors predict long-COVID symptoms. Eur Respir J. 2022.

23. Jia X, Cao S, Lee AS, Manohar M, Sindher SB, Ahuja N, et al. Anti-nucleocapsid antibody levels and pulmonary comorbid conditions are linked to post–COVID-19 syndrome. JCI Insight. 2022;7(13).

24. Giron LB, Peluso MJ, Ding J, Kenny G, Zilberstein NF, Koshy J, et al. Markers of fungal translocation are elevated during post-acute sequelae of SARS-CoV-2 and induce NF-κB signaling. JCI Insight. 2022;7(18).

25. Pretorius E, Venter C, Laubscher GJ, Kotze MJ, Oladejo SO, Watson LR, et al. Prevalence of symptoms, comorbidities, fibrin amyloid microclots and platelet pathology in individuals with Long COVID/Post-Acute Sequelae of COVID-19 (PASC). Cardiovasc Diabetol. 2022;21(1):148.

26. Su Y, Yuan D, Chen DG, Ng RH, Wang K, Choi J, et al. Multiple Early Factors Anticipate Post-Acute COVID-19 Sequelae. Cell. 2022;0(0).

27. Al-Aly Z, Bowe B, Xie Y. Long COVID after breakthrough SARS-CoV-2 infection. Nat Med. 2022:1–7.

28. Xie Y, Choi T, Al-Aly Z. Association of Treatment With Nirmatrelvir and the Risk of Post-COVID-19 Condition. JAMA Intern Med. 2023.

29. Xie Y, Choi T, Al-Aly Z. Molnupiravir and risk of post-acute sequelae of covid-19: cohort study. BMJ. 2023;381:e074572.

30. Durstenfeld MS, Peluso MJ, Lin F, Peyser ND, Isasi C, Carton TW, et al. Association of nirmatrelvir/ritonavir treatment with Long COVID symptoms in an online cohort of non-hospitalized individuals experiencing breakthrough SARS-CoV-2 infection in the omicron era. medRxiv. 2023.

31. Bajema KL, Berry K, Streja E, Rajeevan N, Li Y, Yan L, et al. Effectiveness of COVID-19 treatment with nirmatrelvir-ritonavir or molnupiravir among U.S. Veterans: target trial emulation studies with one-month and six-month outcomes. medRxiv. 2022.

